# Exposure to Higher Cigarette Taxes during Adolescence Reduces Lifetime Smoking in Individuals with Elevated Genetic Risk

**DOI:** 10.1101/2025.09.23.25336460

**Authors:** Robel Alemu, Lauren L. Schmitz

**Affiliations:** University of California Los Angeles (UCLA), 110 Westwood Plaza, Los Angeles, CA 90095, USA; Broad Institute of MIT and Harvard, Program in Medical and Population Genetics, Merkin Building, 415 Main St, Cambridge, MA 02142, USA; Adelaide Medical School, Faculty of Health and Medical Sciences, Adelaide University, Adelaide, Australia; University of Wisconsin-Madison, La Follette School of Public Affairs, 1225 Observatory Dr, Madison, WI 53706

## Abstract

This study examines whether the effects of cigarette taxes on smoking behavior vary by an individual’s genetic predisposition and the timing of policy exposure. Using data from the U.S. Health and Retirement Study, we link historical state-level cigarette excise taxes with polygenic indices (PGIs) for smoking behavior. We find that the timing of tax exposure has distinct effects on different margins of smoking behavior: taxes experienced during adolescence primarily influence long-term smoking participation, while taxes in older adulthood more strongly affect smoking intensity. Our key finding is a significant interaction between adolescent-era taxes and genetic risk. The smoking reduction caused by adolescent taxes is significantly greater for individuals with a higher genetic predisposition to smoke. Conversely, we find no evidence that the effects of taxes in older age are moderated by genetic risk. These results indicate that youth-targeted tax policies are particularly effective for genetically at-risk individuals, and that studies overlooking this heterogeneity may underestimate the long-run benefits of these interventions.

## 1. Introduction

Tobacco use remains a leading cause of preventable death and imposes substantial economic cost in the United States and globally^1–3^. Cigarette taxes are a primary tool for reducing tobacco consumption, yet their effectiveness varies across individuals^4,5^ and their impact on aggregate consumption has recently plateaued^6,7^. This heterogeneity raises a crucial question for economic theory and policy design: what are the mechanisms driving these differential responses, and does the timing of a policy intervention fundamentally alter its long-run impact on population health? A recent literature has begun to explore these questions along two distinct lines of inquiry. One stream of research emphasizes the importance of the timing of policy exposure, demonstrating that higher cigarette taxes experienced during adolescence lead to a significant reduction in smoking and mortality decades later^8^. Concurrently, an emerging literature uses genetic data to show that innate predispositions moderate behavioral response to contemporaneous environmental factors, including older-age cigarette taxes^9,10^. However, it remains unclear whether genetic risk moderates the long-run effects of cigarette taxes experienced during the critical window of adolescence.

This study bridges this gap by providing the first estimates of the interaction between adolescent cigarette taxes and genetic predisposition to smoking. We extend the literature in three important ways. First, we disentangle the effects of cigarette taxes experienced during adolescence from those faced in older age, analyzing their influence across multiple margins of smoking behavior: participation (current smoking), cessation, and intensity (cigarettes per day or CPD). Second, we introduce genetic heterogeneity as a key mechanism explaining differential policy responses, using polygenic indexes (PGIs, also called polygenic scores or genetic risk scores) to test whether the effectiveness of taxes is amplified for genetically predisposed individuals. Third, we use an instrumental variable (IV) approach, leveraging tax-induced variation in smoking to estimate its causal impact on long-run morbidity and mortality.

Researchers are increasingly using PGIs within natural or quasi-natural experiments to assess whether the effects of interventions vary by underlying genetic risk^10–16^. Such advances were made possible in part due to the growing availability of genetic data in large population studies or biobanks^18,19^, and methodological advances in modeling gene-environment interactions (GxE)^12,13,15–17^. PGIs are single-value, objective, time-invariant, continuous indices that summarize an individual’s heritable liability to a phenotype or outcome of interest^20,21^. They are computed as weighted sum of common single nucleotide polymorphisms (SNPs), with each SNP’s allele multiplied by an effect-size weight from genome-wide association study (GWAS) – large studies that tests millions of variant-phenotype associations and are often meta-analyzed across cohorts to improve precision^18,19^. By aggregating small SNP effects, PGIs explain more variation than single SNPs, hence are well suited for assessing the heterogenous effects of policies or interventions^12,20,22^. In the context of smoking, GWAS of smoking behavior have identified hundreds of SNPs linked to smoking initiation, intensity, and cessation ^23–25^. These SNPs are often located in or near genes that regulate nicotine addiction, metabolism, and related neurotransmitter pathways in the brain^25–27^. This genomic evidence aligns with twin and family studies which indicate that smoking behavior is highly heritable (31-60%)^27,28^.

In this study, we exploit plausibly exogenous changes in cigarette taxes across states and over time to assess whether PGIs for smoking behavior moderated the effect of cigarette taxes encountered during adolescence—a critical developmental window in which most individuals initiate smoking—on smoking behavior in adulthood. To conduct our analysis, we use data from the Health and Retirement Study (HRS), which combines rich, population-representative longitudinal data on smoking and health with genetic information for Americans over age 50. This allows us to construct individual histories of tax exposure from adolescence through adulthood and link them to respondents’ PGIs for smoking initiation, cessation, and intensity.

Our results highlight that the timing of tax exposure has notably different effects across the margins of smoking. Adolescent-era taxes are the primary driver of long-run smoking participation and cessation, while taxes experienced during older age are more influential in determining the number of cigarettes consumed by active smokers. Throughout, we use the terms “current smoker” and “lifetime smoker” interchangeably since individuals rarely initiate smoking after age 50, as nearly nine in ten adults in the United States who have ever been daily smokers tried their first cigarette before age 18^29^. Our results show that a one-dollar increase in adolescent-era cigarette taxes is associated with a 5.9 percentage point reduction in the probability of becoming a current or lifetime smoker for an individual with average genetic risk. Notably, this protective effect is significantly larger for those with a higher genetic predisposition; the smoking-reducing effect of the tax increases by 2.7 percentage points for every one standard deviation increase in the smoking initiation PGI. In contrast, the interaction between genetic risk and older-age taxes is small and statistically insignificant for both smoking participation and cessation. The long-run adolescent tax elasticity of smoking participation is −0.41, meaning that a 10% increase in cigarette taxes during adolescence reduces the likelihood of smoking in older adulthood by 4%. This effect is substantially larger in magnitude than the older-age tax elasticity of −0.29.

Finally, our IV analysis reveals that ordinary least squares (OLS) models that do not account for the endogeneity of smoking behavior substantially underestimate the adverse health consequences of smoking. Using the sum of the average cigarette taxes that an individual was exposed to in adolescence and at older ages as an instrument, we find that being a current smoker causally increases the risk of lung disease, heart problems, and cancer. These results underscore the significant long-run welfare gains from tax policies that successfully curb smoking. Taken together, our findings demonstrate that cigarette taxes implemented during the critical window of adolescence have lasting effects that are amplified for genetically at-risk individuals, highlighting the importance of early-life interventions in public health policy.

## 2. Results

### 2.1. Summary statistics

Our final estimation sample includes all genotyped European genetic ancestry HRS respondents with non-missing covariate data (N=11,558). A detailed description and schematic overview of the exclusion criteria are presented in Figure S1. Table S1 summarizes individual, household, and state-level time-invariant and time-varying characteristics for the full sample by smoking status. The average age of respondents was 66.96 (SD=11.01), and about 58 percent were female. A relatively larger share of the respondents (57.02 percent) were ever smokers, meaning they smoked at least 100 cigarettes in their lifetime, 43.29 percent of whom reported quitting smoking at the time of the survey. Due to increases in cigarette taxes over time (Figure S2), the average cigarette excise tax that HRS birth cohorts were exposed to during adolescence ($0.96, SD=0.17) was much lower compared to excise taxes levied when they were surveyed in the HRS after age 50 ($1.52, SD=0.63). Figure S3 depicts the considerable spatial variation in state tobacco excise taxes across states in 2016 (mean=$1.63/ pack, SD=1.16). Current and former smokers have significantly higher PGIs for smoking initiation and intensity than never smokers, which means that, on average, they have more genetic variants that put them at higher risk for becoming smokers and smoking more CPD than never smokers. A standard two-sample t-test in column 6 of Table S1 shows that, compared to never smokers, current smokers had fewer years of education, earned less income, and were more likely to live in states with lower cigarette excise taxes and less stringent tobacco control policies (p<0.01). On average, relative to current or lifetime smokers, former smokers lived longer, were more educated, and resided in states with more stringent tobacco control policies (p<0.01) (column 6 of Table S1). Moreover, former smokers lived in counties with significantly higher travel distances to the nearest lower cigarette excise tax state border than current smokers (p<0.01).

### 2.2. Direct and interactive effects of cigarette taxes and PGIs on smoking behavior

Overall, the PGIs are strong and statistically significant predictors of their corresponding smoking phenotypes (Tables 1-3). As shown in Table S2, the PGI for smoking initiation explains 4% of the variance in current or lifetime smoking, while the PGIs for smoking cessation and CPD explain 2% and 0.56% of the variance in their respective outcomes. The higher predictive power of the smoking initiation PGIs is expected, as the GWAS for this trait can be conducted on much larger samples (including both smokers and non-smokers), whereas the GWAS for cessation and CPD are restricted to smaller samples of individuals who have ever smoked. In comparison to other predictors, the smoking initiation PGI is 14, 3.1, and 1.4 times more predictive than respondents’ age, years of education, and sex, respectively. For smoking cessation and CPD, however, sociodemographic factors such as age and education are more predictive than the corresponding PGIs (Table S2).

**Table 1.** Effect of adolescent and adult cigarette taxes on current smoking at older ages by smoking initiation polygenic index (PGI)

|  | (1) | (2) | (3) | (4) | (5) | (6) |
| --- | --- | --- | --- | --- | --- | --- |
| Adolescent Tax | -0.068***<br>(0.021) | -0.063***<br>(0.021) | -0.066***<br>(0.021) | -0.068***<br>(0.021) | -0.068***<br>(0.021) | -0.059***<br>(0.021) |
| Adult Tax |  | -0.024***<br>(0.008) | -0.025***<br>(0.007) | -0.025***<br>(0.008) | -0.025***<br>(0.008) | -0.026***<br>(0.008) |
| Smoking Initiation PGI |  |  | 0.038***<br>(0.003) | 0.067***<br>(0.014) | 0.069***<br>(0.015) | 0.066***<br>(0.016) |
| Adolescent Tax x PGI |  |  |  | -0.030**<br>(0.013) | -0.029**<br>(0.014) | -0.027*<br>(0.015) |
| Adult Tax x PGI |  |  |  |  | -0.001<br>(0.004) | -0.001<br>(0.005) |
| Min. legal purchase age | No | No | No | No | No | Yes |
| State smoke free air laws | No | No | No | No | No | Yes |
| Travel distance to nearest lower tax state border (level, squared) | No | No | No | No | No | Yes |
| Mean Adolescent Tax |  |  |  |  |  | 0.96 |
| Mean Adult Tax |  |  |  |  |  | 1.52 |
| Mean of dependent variable |  |  |  |  |  | 0.137 |
| Adolescent Tax Elasticity |  |  |  |  |  | -0.413 |
| Adult Tax Elasticity |  |  |  |  |  | -0.288 |
| Individuals (N) | 11,528 | 11,528 | 11,528 | 11,528 | 11,528 | 11,528 |
| Person-wave observations | 110,624 | 110,624 | 110,624 | 110,624 | 110,624 | 110,624 |
| R-squared | 0.095 | 0.095 | 0.107 | 0.108 | 0.108 | 0.109 |

#### 2.2.1. Current smoking

Table 1 presents results for current smoking status at older ages. The models are presented sequentially, demonstrating that our main findings are robust to the inclusion of a comprehensive set of individual and state-level controls. In our preferred specification (column 6), the main effect estimates show that both adolescent and older-age taxes are associated with a lower probability of smoking for an individual with average genetic risk. A one dollar increase in the adolescent-era tax is associated with a 5.9 percentage point reduction in the probability of smoking (p<0.01), while a one-dollar increase in the older-age average tax is associated with a 2.6 percentage point reduction (p<0.01). These effects correspond to a long-run adolescent tax elasticity of smoking participation of -0.41 and an older-age tax elasticity of −0.29. This shows that cigarette taxes experienced during formative years are especially powerful in shaping long-run smoking behavior, with a larger preventive effect than taxes during older adulthood. The magnitude of our main effect for adolescent taxes is larger than that found by Friedson et al.^8^, who in a different sample (Tobacco Use Supplement to the Current Population Survey) and without incorporating genetic risk, report a 1.7 percentage point reduction.

Additionally, we find a significant interaction between adolescent taxes and genetic predisposition. The coefficient on the interaction term (*β*_4_ in equation 4), reported in column 6 of Table 1, is -0.027 (p<0.10), indicating that the protective effect of adolescent taxes is stronger for individuals with a higher genetic risk for smoking. In contrast, the interaction between older-age taxes and the PGI (*β*_5_) is small and not statistically distinguishable from zero.

To illustrate the magnitude of this interaction, we can calculate the total effect of the adolescent tax for different genetic risk profiles. For an individual with a high genetic risk (1 standard deviation above the mean PGI), a one-dollar increase in the adolescent tax is associated with an 8.6 percentage point reduction in the probability of smoking (-0.059 + (−0.027*1)). For an individual with low genetic risk (1 SD below the mean), the same tax increase is associated with a much smaller 3.2 percentage point reduction (-0.059 + (−0.027*-1). This demonstrates that the long-term deterrent effect of taxes experienced during the critical window of adolescence is significantly amplified for those who are most genetically predisposed to smoking.

Our finding of a null interaction for average taxes at older ages differs from Slob and Rietveld^10^, who, using the same HRS data, report a significant PGI-by-tax interaction of -0.012 for current smoking. However, direct comparison is difficult as their model does not control for adolescent taxes and uses a different measure of older-age taxes. The null interaction in our model for adult taxes suggests that after accounting for adolescent taxes, the effect of taxes on current smoking after age 50 does not vary by genetic risk.

#### 2.2.2. Smoking cessation

Table 2 presents the results for smoking cessation for former versus current smokers. In our preferred specification (column 6), we find that older-age taxes are associated with smoking cessation. A one dollar increase in adult taxes is associated with a 2.7 percentage point increase in the probability of being a former smoker (p<0.05). In contrast, the main effect of adolescent taxes on cessation is positive but not statistically significant. The corresponding tax elasticities of cessation are 0.081 for adolescent taxes and 0.055 for older-age taxes.

**Table 2.** Effects of adolescent and adult cigarette taxes on smoking cessation by smoking cessation polygenic index (PGI)

|  | (1) | (2) | (3) | (4) | (5) | (6) |
| --- | --- | --- | --- | --- | --- | --- |
| Adolescent Tax | 0.089**<br>(0.043) | 0.082*<br>(0.043) | 0.078*<br>(0.043) | 0.080*<br>(0.043) | 0.080*<br>(0.043) | 0.065<br>(0.041) |
| Adult Tax |  | 0.025**<br>(0.012) | 0.025**<br>(0.012) | 0.025**<br>(0.012) | 0.025**<br>(0.012) | 0.027**<br>(0.014) |
| Smoking Cessation PGI |  |  | -0.029***<br>(0.006) | -0.065***<br>(0.020) | -0.058***<br>(0.021) | -0.063***<br>(0.021) |
| Adolescent Tax x PGI |  |  |  | 0.037<br>(0.023) | 0.040<br>(0.024) | 0.042*<br>(0.024) |
| Adult Tax x PGI |  |  |  |  | -0.006<br>(0.007) | -0.006<br>(0.007) |
| Min. legal purchase age | No | No | No | No | No | Yes |
| State smoke free air laws | No | No | No | No | No | Yes |
| Travel distance to nearest lower tax state border (level, squared) | No | No | No | No | No | Yes |
| Mean Adolescent Tax |  |  |  |  |  | 0.96 |
| Mean Adult Tax |  |  |  |  |  | 1.52 |
| Mean of dependent variable |  |  |  |  |  | 0.137 |
| Adolescent Tax Elasticity |  |  |  |  |  | -0.413 |
| Adult Tax Elasticity |  |  |  |  |  | -0.288 |
| Individuals (N) | 6,587 | 6,587 | 6,587 | 6,587 | 6,587 | 6,587 |
| Person-wave observations | 62,668 | 62,668 | 62,668 | 62,668 | 62,668 | 62,668 |
| R-squared | 0.141 | 0.141 | 0.144 | 0.144 | 0.144 | 0.146 |

*Notes:* This table presents coefficients from linear probability models that pool observations across all waves, based on equation (4). The dependent variable is an indicator for whether a respondent is a former smoker. Adolescent Tax is the average state cigarette excise tax experienced during adolescence, and Adult Tax is the average state tax experienced during older adulthood. PGI is the standardized polygenic index for smoking cessation. All models control for sex, a quadratic in age, a quadratic in years of education, marital status indicators, and annual household income quintiles. All models also include fixed effects for birth year, adolescent state of residence, current state of residence, survey year, a current-state-specific linear time trend, and the first 10 PCs of the genotype data. State-level controls include smoke-free air laws, minimum legal purchase age, and the unemployment rate. Robust standard errors, shown in parentheses, are two-way clustered at the individual and adolescent state-of-residence levels. The tax elasticities in Model 6 are calculated at the sample mean as follows: Adolescent Tax Elasticity = $(0.065 + (0.042 * 0)) * (0.95 / 0.759) \approx 0.081$ ; Adult Tax Elasticity = $(0.027 + (-0.006 * 0)) * (1.51 / 0.759) \approx 0.055$ . \*\*\* $p < 0.01$ , \*\* $p < 0.05$ , \* $p < 0.1$ .

Turning to the interaction effects, we find no evidence that the effect of adult taxes on quitting varies by genetic predisposition. Our finding of null interaction for older-age taxes is consistent with Slob and Rietveld^30^, who also do not find a significant genetic gradient in the effect of older-age taxes on cessation, although their models do not account for adolescent taxes. However, we find a marginally significant interaction between adolescent taxes and the smoking cessation PGI (p<0.10). While the main effect of adolescent taxes is not significant for the average-PGI individual, the significant interaction term indicates that the total effect of the tax does vary by genetic risk. The positive coefficient suggests that higher adolescent taxes were more effective in promoting cessation among individuals with a higher genetic predisposition to continue smoking. Specifically, for every one standard deviation increase in the PGI, the quitting-promoting effect of a one dollar adolescent tax increases by 4.2 percentage points.

#### 2.2.3. Smoking Intensity

Table 3 presents the results for smoking intensity, measured as the natural log of cigarettes per day (CPD), for the subsample of current smokers. In our full specification (column 6), we find that smoking intensity is responsive to taxes experienced in older age, but not to those experienced during adolescence. The coefficient on the log of adult taxes is −0.286 (p<0.01). In the log-log specification, this coefficient is interpreted as the tax elasticity of smoking intensity, indicating that a 10% increase in the older-age tax is associated with a 2.86 decrease in the number of cigarettes smoked per day. The magnitude of this elasticity is larger than the -0.03 to -0.04 tax elasticity range found by MacLean et al.^31^ using the same HRS data; however, their study uses survey-year specific cigarette tax exposure while we use the average tax exposure during older adulthood, and they do not account for genetic risk.

**Table 3.**
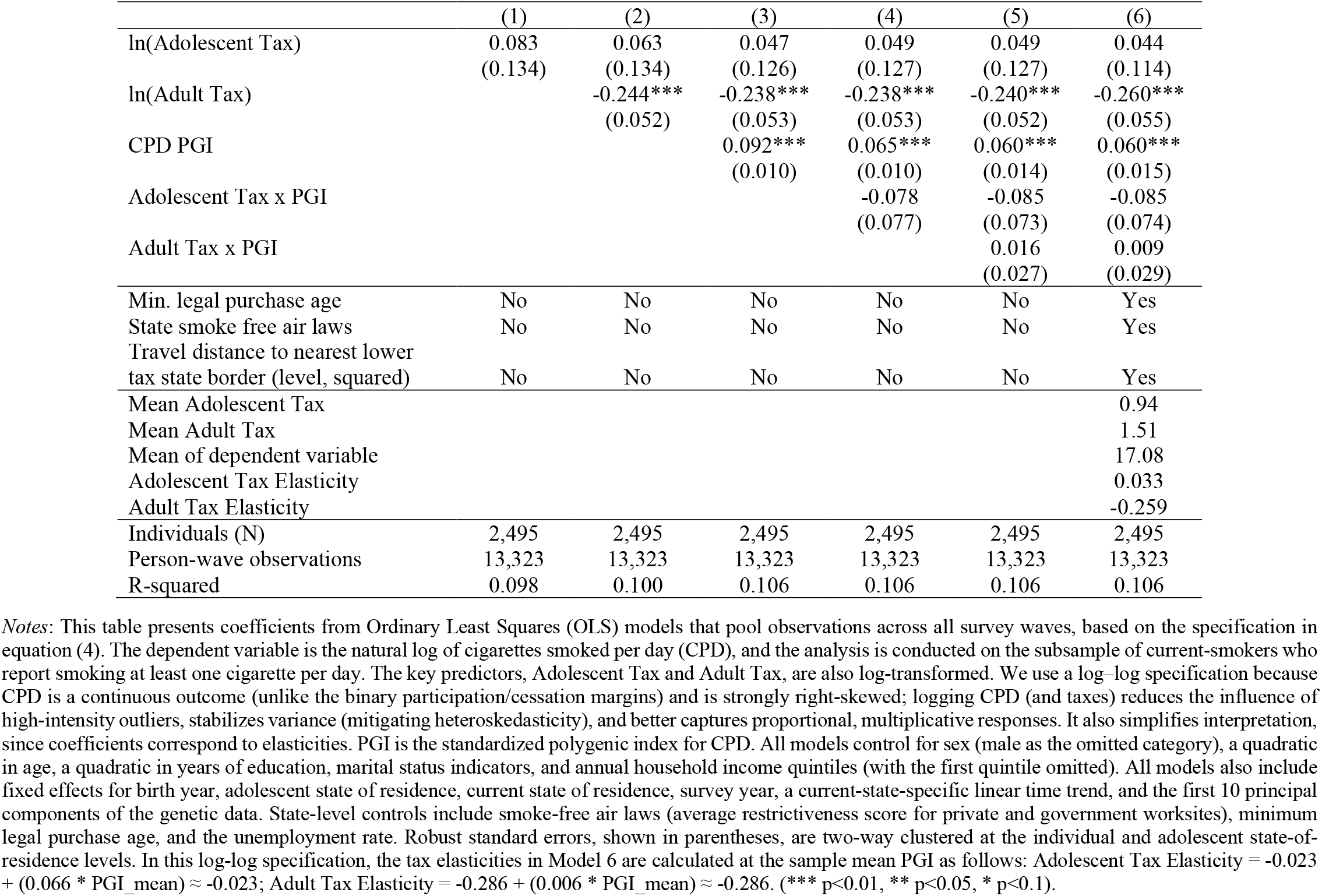
Effects of adolescent and adult cigarette taxes on cigarettes smoked per day (CPD) by CPD polygenic index (PGI)

We do not find evidence that the effects of either adolescent or adult taxes on CPD vary by an individual’s genetic predisposition, as both interaction terms are small and statistically insignificant. Our null finding for older-age taxes differs from Slob and Rietveld^10^, who find a marginally significant genetic gradient (p<0.10); however, their model does not account for adolescent taxes, which likely explains the difference in results. This overall pattern suggests that while adolescent-era taxes are crucial in shaping the decision to smoke at all (the extensive margin), the quantity of cigarettes consumed by active smokers is more sensitive to contemporaneous tax rates. It is also important to note that these models may be underpowered, as they are run on a subsample of smokers who report smoking at least one cigarette per day, which comprises only 22% (2,533 out of 11,528) of the full analysis sample.

A more intuitive understanding of the genetic gradient in the effects of cigarette taxes on smoking can be gleaned from the two-way contour plots in Figures 1, S4, and S5. The plots depict the predicted probabilities of smoking across PGI values and cigarette excise tax exposure. The patterns in Figure 1 indicate that higher genetic risk individuals were more likely to become lifetime or current smokers when exposed to lower cigarette taxes than those experiencing higher cigarette taxes. Figure 2 further simplifies the interpretation of the observed genetic gradient by plotting coefficients from a model where the PGI is categorized into three groups: Low (bottom 20%), Medium (middle 60%), and High (top 20%). Relative to the Medium PGI reference group, the interaction for adolescent taxes (Panel A) is positive and statistically significant for the High PGI group, indicating a stronger smoking-reducing effect of the tax for this group. Conversely, the interaction for older-age taxes (Panel B) is not statistically significant for any group. This categorical analysis reinforces our primary finding from the continuous interaction model: the moderating effect of genetic risk is unique to taxes experienced during adolescence. Furthermore, a genetic risk gradient also emerges in the effects of cigarette taxes on the likelihood of quitting (Figure S4). Specifically, panel A of Figure S4 reveals that those with a higher genetic propensity to avoid quitting were more responsive to marginally higher adolescent cigarette taxes than those with a lower genetic predisposition to resist quitting.

**Figure 1.**
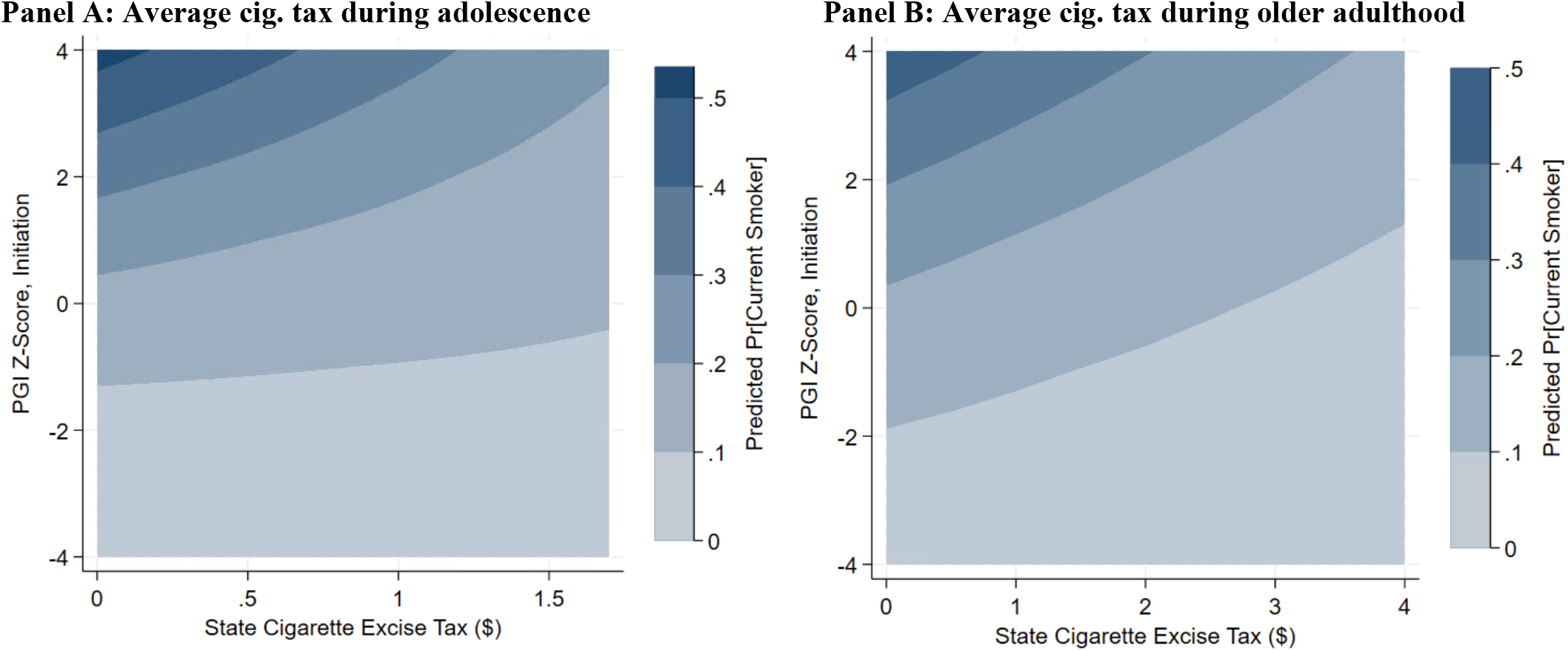
Predicted probability of becoming a current smoker by PGI and life course timing of cigarette excise taxes. *Notes*: These contour plots show the predicted probability of becoming a current smoker for respondents who experienced different levels of average cigarette excise tax during their adolescence (panel A) and while in the HRS (panel B) by genetic predisposition to smoking initiation (PGI Z-score). The 3^rd^ axis (z-axis) of the contour plot depicts the marginal predicted probabilities, which were estimated using a logistic regression model run on the HRS dataset that pools observations across all survey waves as per equation (4). Contours of similar color represent identical predicted probabilities of being a lifetime smoker.

**Figure 2.**
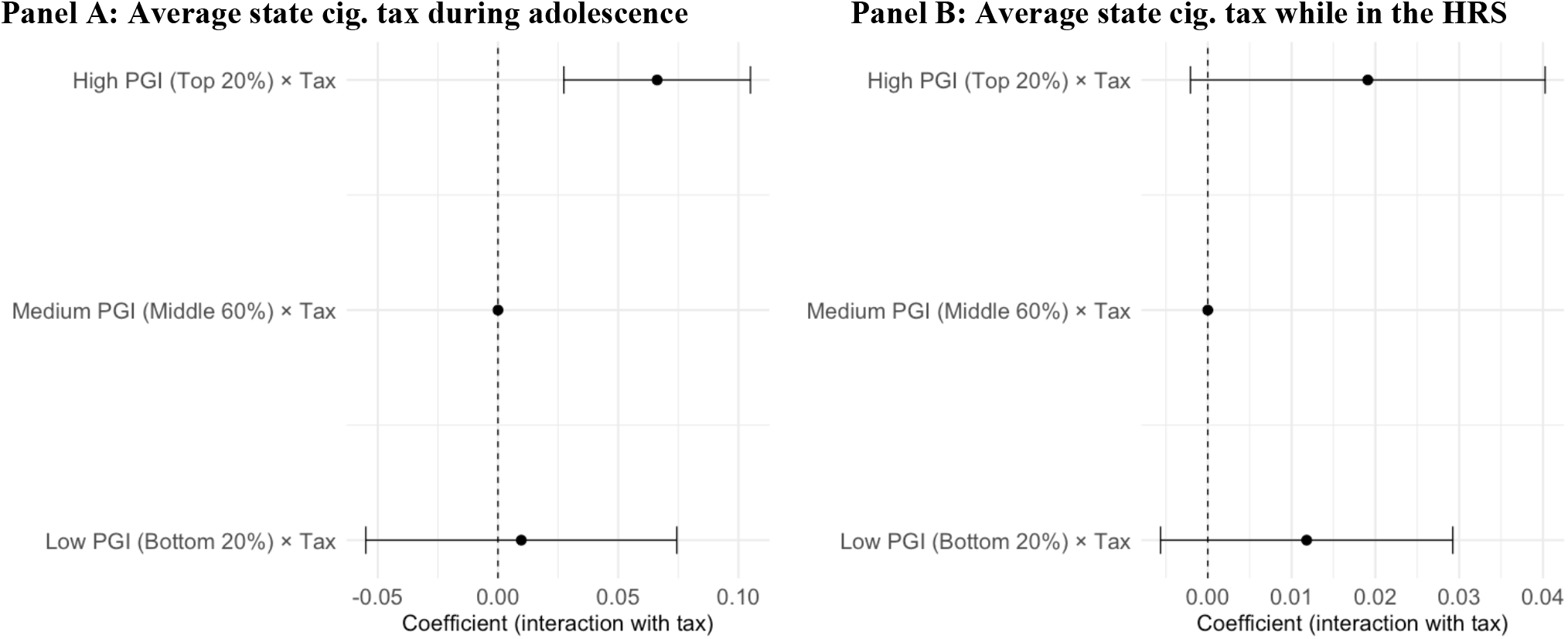
Effects of cigarette taxes on the likelihood of being a lifetime smoker by genetic risk profile category. *Notes*: Panels A and B present estimates from regression models similar to those in equation (4), except the continuous smoking initiation polygenic index (PGI) is replaced with a categorical version. The PGI is divided into three mutually exclusive groups: Low PGI (bottom 20% of the distribution), Medium PGI (middle 60%), and High PGI (top 20%). The Medium PGI group serves as the reference category and is shown as a point at zero without a confidence interval. Black dots represent the estimated interaction coefficients between cigarette excise taxes and the Low or High PGI categories relative to the Medium group, with horizontal lines indicating 95% confidence intervals. Panel A uses the average state cigarette excise tax respondents faced during their teenage years (“adolescent taxes”), and Panel B uses the annual state cigarette excise tax at the time respondents were observed at older ages. The dependent variable in both panels is an indicator for whether respondents are lifetime smokers.

### 2.3. Effects of cigarette-tax-induced changes in smoking on health

Our core results show that individuals with higher PGIs for smoking tend to benefit from higher cigarette excise taxes, as it increases the likelihood of smoking cessation, above and beyond what is expected for individuals with average PGIs. Having detected a robust PGI-by-tax interaction effect, we now assess how changes in smoking behavior induced by higher cigarette taxes impacted the likelihood of developing chronic illnesses and survival. To realize this, we employ an instrumental variable (IV) approach to estimate the causal link between smoking and various health indicators. We present coefficients from two-stage least squares (2SLS) models relative to naïve OLS models in Table 4 and Tables S4-S5. Our instrument, the sum of average cigarette excise taxes experienced during adolescence and older age, meet the relevance assumption of 2SLS models as all of our 1^st^ stage regressions had high F-statistics greater than 10, the rule of thumb threshold for 2SLS models.

**Table 4.** Effects of being a current smoker on the likelihood of developing chronic diseases and on survival.

|  | Lung Disease |  | Heart Problem |  | Any Cancer |  | Diabetic |  | Survival |  |
| --- | --- | --- | --- | --- | --- | --- | --- | --- | --- | --- |
|  | (1) | (2) | (3) | (4) | (5) | (6) | (7) | (8) | (9) | (10) |
|  | OLS | 2SLS | OLS | 2SLS | OLS | OLS | OLS | 2SLS | OLS | 2SLS |
| Current Smoker | 0.101***<br>(0.006) | 0.680***<br>(0.143) | 0.005<br>(0.009) | 1.226***<br>(0.255) | 0.001<br>(0.008) | 0.571***<br>(0.169) | -0.000<br>(0.007) | 0.661***<br>(0.160) | -0.021***<br>(0.003) | -1.525***<br>(0.270) |
| F-stat (1<sup>st</sup> Stage) | - | 37.59 | - | 37.84 | - | 36.40 | - | 38.37 | - | 43.37 |
| Per-wave observations | 97,487 | 97,487 | 97,487 | 97,487 | 97,487 | 97,487 | 97,487 | 97,487 | 105,067 | 105,067 |
| Individuals (N) | 11,703 | 11,703 | 11,703 | 11,703 | 11,703 | 11,703 | 11,703 | 11,703 | 11,703 | 11,703 |

Notes: This table shows coefficients of naïve OLS (models 1, 3, 5, 7, and 9) and 2SLS (models 2, 4, 6, 8, and 10) regression models based on the specifications highlighted in equations (6-9). The dependent variables are indicators for whether respondents report ever being diagnosed with lung disease (models 1 and 2), heart problems (models 3 and 4), any cancer (models 5 and 6), diabetes (models 7 and 8), and survival at the time of survey (models 9 and 10). Models 1-8 were run on the HRS dataset that pools observations across all survey waves (1992-2016), conditional on survival at a given survey wave. The key predictor is an indicator variable for whether a respondent is a "Current smoker". In the 2SLS models, smoking behavior or "Current Smoker" is instrumented by the average state cigarette excise tax that respondents were exposed during adolescence and while in the HRS. All regression models control for gender, BMI (level, squared), education (level, squared), marital status, household income, state, birth year, and survey-year fixed effects. Robust standard errors clustered at the individual and state of residence levels are shown in parentheses. \*\*\* p<0.01, \*\* p<0.05, \* p<0.1.

Overall, we find that the endogenous OLS model consistently underestimates the effects of smoking (both in the extensive and intensive margins) on several chronic illnesses and survival. In Table 4, we show that being a lifetime smoker increases the risk of developing lung disease, heart problems, cancer (any), diabetes, and death at the time of the survey (p<0.01). Our 2SLS estimates, for instance, show that being a current smoker increases the likelihood of reporting any form of lung disease by 68 percent, while the OLS indicates an increase of just 10.1 percent. Similarly, the 2SLS models show that being a current smoker increases the risk of reporting any heart problem (123%), any form of cancer (57.1%), and diabetes (66.1%), whereas the effects were null in the OLS models. Furthermore, being a lifetime smoker decreases survival at the time of the survey by 152.5 percent based on our 2SLS model, while the decline in survival was just 2 percent in the OLS model. In addition, we also show the impacts of other smoking phenotypes (quitting and CPD) in our supplementary information (Tables S4 and S5). Overall, we find qualitatively similar effects of smoking on the health outcomes in question, and that the OLS models are consistently downward-biased.

### 2.4. Robustness analyses

Our main analysis is restricted to HRS participants who survived until at least 2006 to provide genetic data. As shown by Domingue et al.^32^, this can introduce mortality selection bias, as survivors tend to be healthier and less likely to smoke than those who died before 2006. To assess the sensitivity of our estimates to this potential bias, we re-estimate our main specification using an inverse probability weighting (IPW) approach to account for differential survival (see Supplementary Note A for details). We find that our results are robust to this correction. After applying the weights, the magnitude of the key interaction term for adolescent taxes and the smoking initiation PGI on current smoking is consistent, confirming that our main findings are not driven by mortality selection (Table 5).

**Table 5.** Effects of cigarette taxes on current smoking by smoking initiation PGI (adjusted for survival selection)

|  |  |
| --- | --- |
| Adolescent Tax | -0.048**<br>(0.022) |
| Adult Tax | -0.046**<br>(0.014) |
| Smoking Initiation PGI | 0.076***<br>(0.018) |
| Adolescent Tax x PGI | -0.029*<br>(0.017) |
| Adult Tax x PGI | -0.008<br>(0.006) |
| Per-wave observations | 96,996 |
| Individuals (N) | 9,719 |
| R-squared | 0.118 |

*Notes:* This table presents coefficients from a linear probability model for current smoking status. The model is identical to the main specification (Table 1, Model 6) but is weighted using inverse probability weights (IPWs) to correct for mortality selection based on survival to 2006. The dependent variable is an indicator for whether a respondent is a current smoker. Adolescent Tax is the average state cigarette excise tax experienced during adolescence, and Adult Tax is the average state tax experienced during older adulthood. PGI is the standardized polygenic index for smoking initiation. The model includes the full set of individual, state-level, and genetic controls, as well as all fixed effects detailed in the notes for Table 1. Robust standard errors, shown in parentheses, are two-way clustered at the individual and adolescent state-of-residence levels. \*\*\* p<0.01, \*\* p<0.05, \* p<0.1.

Second, following Keller^33^, we address potential bias that can arise when population structure correlates with environmental exposures (e.g., state cigarette taxes) by including interactions between each of the top ten genetic principal components (PCs) and both the adolescent and adult tax variables. This specification allows tax effects to vary along ancestry-related axes of genetic variation, reducing the risk that ancestry-correlated environmental structure is misattributed to the PGI. Estimates from this specification (Supplementary Table S3) are qualitatively unchanged relative to the main results: the adolescent tax main effect and its interaction with the smoking-initiation PGI remain negative and statistically significant, while the corresponding adult-tax interaction remains small and imprecise. Together with the IPW results above, these findings indicate that our core conclusions are robust to potential biases related to selective survival and population structure^33^.

## 3. Discussion

In this study, we examine how the long-run effects of cigarette taxes vary by genetic predisposition and the timing of policy exposure. Our results yield two primary findings. First, we show that the timing of tax exposure has distinct effects on different margins of smoking behavior. Taxes experienced during adolescence are the most powerful determinants of long-run smoking status, while taxes experienced in adulthood are associated with smoking cessation and smoking intensity among active smokers. Second, we provide evidence that the long-run deterrent effect of adolescent-era taxes on current smoking is significantly amplified for individuals with a higher genetic predisposition to smoke. In contrast, we find no evidence that the effects of older-age taxes on either smoking participation or intensity are moderated by genetic risk.

Our results have broader implications for both economic theory and public policy. Regarding theory, the canonical rational addiction model^34^ in economics provides a framework for understanding addictive behaviors by assuming that individuals make forward-looking decisions, weighing current consumption against expected future costs. However, this model does not explicitly incorporate innate predisposition to addiction. Later extensions allow for stable individual differences – such as variation in how quickly the harmful effects of past smoking accumulate (the addictive stock), how heavily individuals value immediate gratification relative to long-term health (discounting behavior), or how sensitive they are to the reinforcing effects of consumption^35–39^. By using a PGI as a direct proxy for such predispositions – rather than treating it as an unobserved parameter – we can examine heterogeneity in tax responsiveness along a dimension that the original model could not measure directly. The significant interaction between adolescent cigarette taxes and genetic risk suggests that early-life price shocks are particularly effective for those with a high genetic predisposition to smoke, which is in line with the predictions of the rational addiction model. This result would be obscured in analyses that ignore genetic heterogeneity^8,31^. From a policy perspective, this underscores that youth-targeted tobacco taxes are not only broadly effective but also disproportionately benefit those most at risk of initiating smoking. Moreover, our findings suggest that the recently observed stagnation in cigarette tax responsiveness may be temporary and could reverse as newer cohorts of high-risk individuals are exposed to progressively higher taxes during adolescence^8^.

Beyond smoking behavior, our analysis demonstrates that the reductions in smoking attributable to higher cigarette taxes are associated with substantial long-term health benefits. Using an IV 2SLS approach to address endogeneity, we find that OLS models substantially understate the adverse effects of smoking on chronic disease and survival. The 2SLS estimates indicate large causal impacts of smoking on lung disease, heart problems, cancer, diabetes, and mortality, highlighting that policies which effectively reduce smoking—whether by preventing initiation or promoting cessation—can yield significant gains in population health. These findings underscore the broader welfare implications of tobacco tax interventions.

Certain aspects of our study design point to important avenues for future research. First, our analysis is restricted to respondents of European genetic ancestry, meaning our findings may not generalize to other genetic ancestries. As genetic data from more diverse samples become available, future research will be crucial to evaluate the replicability of these findings across different genetic ancestries. Second, the PGIs used in this study are derived from standard (between-family) GWAS. Studies have shown that such PGIs are susceptible to biases from factors such as gene-environment correlation (including indirect genetic effects and population stratification) and assortative mating, which could, in turn, bias our estimates^40,41^. Future research could test the validity of the genetic gradient found here by using PGIs derived from within-family GWAS design, which mitigate such biases by more cleanly isolating direct genetic effects^40,42,43^. Despite these considerations, this study provides novel evidence on the interplay between the timing of policy interventions and innate genetic risk, highlighting the long-run benefits of early-life policy interventions.

## 4. Data and Methods

### 4.1. Description of study cohort

The Health and Retirement Study (HRS) is a nationally representative longitudinal study of individuals aged 50 plus and their spouses in the U.S. We use the RAND Longitudinal File, which contains harmonized data from 13 waves of HRS interviews (1992-2018)^44,45^. This dataset provides comprehensive information on respondents’ socioeconomic status, demographic characteristics, health behaviors, and physical/functional ability. In addition, we use restricted geographic data to identify respondents’ state of residence during primary schooling years (adolescence) and while in the HRS (adulthood)^44,45^. The initial sample from these waves includes 42,719 individuals. From this sample, we apply several restrictions to arrive at our final estimation sample (see Figure S1 for a detailed description). First, we restrict the analysis to individuals of European genetic ancestry, as the PGIs used in this study were derived from SNP weights that were estimated in individuals of European ancestry and hence do not predict phenotypes as accurately in other genetic ancestries. The sample is further limited by excluding respondents who were deceased prior to the first wave of genotyping in 2006, as well as those who were alive in 2006 or in subsequent waves but were not genotyped. Finally, we exclude individuals with missing values for any covariates used in our regression models. These criteria yielded a final estimation sample of 11,558 individuals (Figure S1).

### 4.2. Assessment of smoking behavior

From the HRS survey waves, we construct three measures of smoking behavior: current smoking, smoking cessation, and smoking intensity. We create an indicator for whether a respondent currently smokes (Current Smoker) and an indicator for smoking cessation (Former Smoker). Given the low probability of initiating smoking after age 50, we also classify current smokers in the HRS as “lifetime smokers.” For smoking intensity, we construct a continuous variable for the number of cigarettes smoked per day (CPD) among current smokers. If responses were reported in packs per day they were converted to cigarettes (1 pack = 20 cigarettes). The use of maximum CPD for former smokers is supported by prior work that found a strong correlation between average and maximum CPD over time^46^. While these self-reported measures do not capture all aspects of tobacco use, such as depth of inhalation or mode of consumption, they provide a reasonable proxy for consumption^47–49^.

### 4.3. Measuring genetic risk for smoking

PGIs were constructed for each respondent by summing the number of risk-associated alleles they carry at each SNP, weighted by the strength of the SNP’s association with the smoking trait of interest estimated from a GWAS^50^. Specifically, SNP weights are the effect-size estimates from a regression of the trait on each SNP, run one at a time:

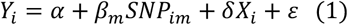

where *Y*_*i*_ is the smoking trait for individual *i*, and *SNP*_*im*_ is the genotype of individual *i* at locus *m*, coded as 0, 1, or 2 depending on the number of reference alleles present. The PGI for each individual is then constructed as the weighted sum of their genotypes across all M SNPs included in the index:

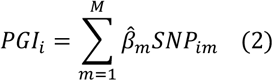

PGIs constructed from the most recent GWAS explain approximately 5-10% of the variance in smoking behavior, net of the variance explained by standard covariates^24,51,52^. The PGIs used in this study were constructed in the HRS using GWAS summary statistics from discovery samples that exclude HRS. For smoking initiation and cessation, we use PGIs developed by the Social Science Genetic Association Consortium (SSGAC)^51^, which have superior predictive accuracy as they were generated by meta-analyzing summary statistics from several large-scale GWASs, including the UK Biobank and 23andMe. As a GWAS for current smoking is not yet available, we use the smoking initiation PGI in our analysis of current smoking status. For smoking intensity (CPD), we rely on the CPD PGI released by the HRS, as the SSGAC did not generate smoking intensity PGIs^53^. For ease of interpretation, all PGIs are standardized to have a mean of zero and a standard deviation of one. Higher PGIs for smoking initiation, cessation, and CPD are associated with worse outcomes (i.e., a higher probability of becoming a regular smoker, a higher probability of not quitting, and increases in CPD). A critical consideration when using PGIs is that GWAS are often performed within a specific genetic ancestry as differences in allele frequencies and linkage disequilibrium (the correlation structure between SNPs) can distort estimated relationship in pooled samples^54–56^. To date, genotyped sample sizes for populations of non-European genetic ancestries have not been large enough to produce well-powered GWAS for smoking behavior^24^. Consequently, we restrict our analysis to individuals of European genetic ancestry, as the available PGIs do not have the same predictive power for individuals from other genetic ancestries, and comparisons across groups could confound genetic effects with environmental differences^57^. Even within our European genetic ancestry sample, we must account for population stratification – the possibility that subtle genetic structure arising from non-random mating could be spuriously correlated with our outcomes of interest^58,59^. To address this, we control for the first 10 principal components (PCs) of the genotype data in all regression models, a standard approach to account for fine-scale population structure^59–61^.

### 4.4. Measuring exposure to cigarette taxes

Data on annual state-level excise tax per cigarette pack comes from the historical Tax Burden on Tobacco dataset^62^. The data spans all years that HRS respondents were in adolescence and/or adulthood (1940-2018) (Figures S2 and S4). We adjust for inflation using the Consumer Price Index with 1991 as the base year. To measure cigarette tax exposure during adolescence, we calculate the average per-pack cigarette tax levied in the respondent’s state of residence when they were between 10 and 18 years of age. Specifically, the HRS asks respondents what state they lived in most of the time when they were in grade school, high school, or around age 10. Since this is the only information available on state of residence after birth until individuals are surveyed in the HRS after age 50, we assume the same state of residence for ages 10-18. Here, we closely following the setup employed by Friedson et al.^8^:

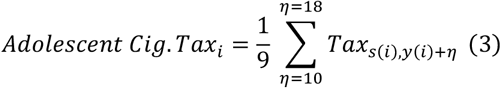

where *y(i)* is respondent *i*’s year of birth and *s(i)* is their state of residence during adolescence. Cigarette taxes at older ages (i.e., after age 50) were linked to respondents based on their current state of residence at each wave. Similarly, tax exposure at older ages was averaged across waves in which respondents were observed in the HRS as per equation (3).

### 4.5. Accounting for other tobacco control policies

It is plausible that certain time-varying factors across states, such as anti-smoking sentiment, could be correlated with both cigarette taxes and smoking behavior, biasing the estimated tax effects^63^. To address this concern, we control for a comprehensive set of state-level policies and economic conditions. We compile data on smoke-free air (SFA) laws from the ImpacTeen Project and the Americans for Non-Smokers’ Rights Foundation (ANRF)^64^, from which we construct a variable averaging the restrictiveness score of bans across private and government worksites. These restrictiveness scores range from 0 (no ban) to either 3 or 5, with higher scores indicating more comprehensive and restrictive bans. For each respondent, we calculate the average score across all survey years to reflect cumulative exposure. We also source data on the minimum legal purchase age (MLPA) from the ImpacTeen Project (ImpacTeen.org, 2009). Finally, we include the annual state unemployment rate from the Bureau of Labor Statistics to account for economic conditions. All state-level datasets were merged with the HRS data by state and survey year.

### 4.6. Proxy measure for cigarette tax avoidance behavior

A potential source of bias in estimating the effects of cigarette taxes is tax avoidance behavior, as price-sensitive smokers may purchase cigarettes from neighboring states with lower taxes^63,65^. To address this, our models control for the opportunity to engage in cross-border cigarette purchases. We construct a proxy for this behavior by computing the travel distance in miles from the centroid of a respondent’s county of residence in adulthood to the border of the closest state with a lower cigarette tax (level and squared). This calculation was performed for each respondent in each survey wave using geospatial analysis tools in ArcGIS Pro (Esri, v3.2, Redlands, CA, 2024) (details in Supplementary Note B).

### 4.7. Other covariates

Our regression models include a rich set of individual-level control variables to account for demographic and socioeconomic factors correlated with smoking behavior. Demographic controls include an indicator for sex (female, with male as the omitted category) and a quadratic for age. Socioeconomic status is controlled for using a quadratic for years of education and a series of indicator variables for marital status (divorced, separated, widowed, or never married, with married or partnered as the reference category). To control for household income, we convert annual household income to 2008 dollars using the CPI and construct a set of five indicators for income quintiles, with the first quintile as the omitted category.

### 4.8. Empirical Strategy

#### 4.8.1. Testing for effects of cigarette taxes on smoking by genetic predisposition

Our empirical strategy is designed to test whether the demand response to adolescent-era cigarette taxes varies by an individual’s genetic predisposition, while disentangling these effects from taxes faced in older adulthood. We analyze smoking behavior across three margins: current smoking and smoking cessation (extensive margins), and smoking intensity (as measured by CPD, intensive margin). Our primary analysis uses a pooled cross-section of all surveys, closely following the model specifications of prior literature^8,31^. The rationale for pooling observations is that there is little within-individual variation in smoking behavior for this older cohort. For our binary outcomes, we estimate a linear probability model (LPM), and for our continuous outcome, we run an ordinary least squares (OLS) model, both within a high-dimensional fixed-effects framework.

Our general specification is as follows:

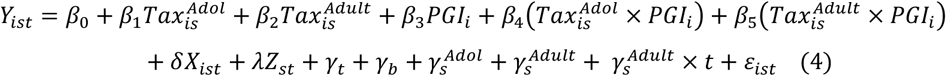

This equation is adapted for each margin of smoking. For the extensive margin, *Y*_*ist*_ is a binary indicator for current or former smoker; 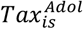 is the average annual cigarette excise tax that the HRS respondent experienced during adolescence, and 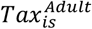 is the average annual cigarette tax that respondents were exposed to while in the HRS; *PGI*_*i*_ is the individual’s PGI for the respective smoking outcome of interest (i.e., smoking initiation, cessation, or CPD). For current smoking and cessation, the coefficients *β*_1_ and *β*_2_ are interpreted as the percentage point change in the probability of the outcome for a one dollar increase in the adolescent or adult tax, respectively. For the intensive margin, *Y*_*ist*_ is the natural log of CPD and the tax variables are also log-transformed. In this log-log specification, the coefficients *β*_1_ and *β*_2_ are interpreted as tax elasticities. Current smoking models are estimated in the sample of current, former, and never smokers; cessation models are estimated in the subsample of current and former smokers; CPD models were estimated in the subsample of current smokers.

The primary coefficients of interest are *β*_4_ and *β*_5_, which capture how the effects of adolescent and older-adulthood taxes are moderated by genetic risk. A negative value for *β*_4_ in the current smoking model would indicate that the smoking-reducing effect of adolescent taxes is stronger for individuals with a higher genetic predisposition for smoking initiation.

In all specifications, we include a vector of individual-level controls *X*_*ist*_ (female, age, age squared, years of education and its square, marital status, household income, and the first 10 PCs of the genotype data), and a vector of time-varying state-level controls for current state of residence *Z*_*st*_ (unemployment rate, smoke-free air laws, and a proxy for cigarette tax avoidance). To control for unobserved heterogeneity, the model includes a full set of fixed effects for survey year (*γ*_*t*_), birth-year (*γ*_*b*_), state of residence during adolescence 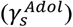, and current state of residence 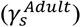. The geographic fixed effects help absorb time-invariant differences at the state level, while the time fixed effects absorb factors that vary over time but are invariant across states. Furthermore, to account for smooth, unobserved changes within states over time, we also include state-specific linear time trends 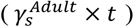. *ε*_*ist*_ is the idiosyncratic error term. For our main pooled models, standard errors are two-way clustered at the individual level to correct for autocorrelation across survey waves for the same person, and at the adolescent state-of-residence level to account for correlated shocks among individuals who grew up in the same state.

Finally, we compute tax elasticities for all outcomes. In this context, elasticities express the percentage change in smoking behavior associated with a 1% change in cigarette taxes, making them a standard metric in health economics and public health for comparing the effectiveness of price-based interventions across settings and populations. For the log-log CPD model, the coefficients on the logged tax variables (adjusted for the interaction term) are the elasticities. For the LPM on current smoking and cessation, we calculate the elasticities at the sample mean using the standard formula 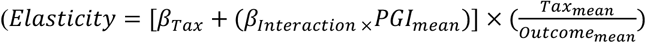.

#### 4.8.2. Estimating the causal effects of smoking on health

Our second objective is to estimate the causal impact of smoking on long-run health outcomes. We first estimate a naïve OLS model, which regresses health directly on smoking behavior, as a baseline for comparison:

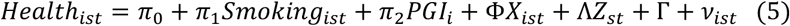

This model is likely biased due to the endogeneity of smoking behavior, which can arise from two primary sources. First, there may be unobserved confounding variables, such as an individual’s risk preference or discount rate, that are correlated with both the decision to smoke and other health-related behaviors. For instance, individuals with a higher tolerance for risk may be more likely to smoke and less likely to engage in preventive healthcare, leading OLS to overstate the causal effect of smoking on poor health. Second, reverse causality is a significant concern; a negative health shock, such as a lung disease diagnosis, may cause an individual to quit or reduce their smoking. This would lead OLS to underestimate the true adverse effect of smoking, as the illness itself reduces the observed prevalence of smoking in the sample.

To address this, we use an instrumental variable (IV) approach with a two-stage least squares (2SLS) estimator. We instrument for an individual’s smoking behavior – across all three margins of smoking – using their average lifetime exposure to state-level cigarette taxes. Specifically, our instrument is the sum of the average cigarette excise tax a respondent was exposed to during adolescence and the average tax they were exposed to during older adulthood (i.e., while in the HRS). Cigarettes are a valid instrument as they strongly predict smoking behavior (relevance) but are plausibly uncorrelated with an individual’s health outcomes except through their effect on smoking (exclusion restriction)^8,66^.

The first stage of our 2SLS model regresses the endogenous smoking variables on the tax instrument and the full set of exogenous controls:

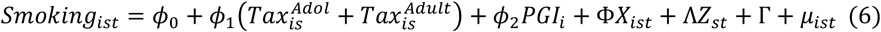

where *Smoking*_*ist*_ is the measure of smoking behavior (current smoking, cessation and CPD); the model includes the same vector of individual controls (*X*_*ist*_), state-level controls (*Z*_*st*_), and the full set of fixed effects (Γ as in equation (3).

The second stage uses the predicted value of smoking from the first stage to estimate the average causal effect of smoking on health:

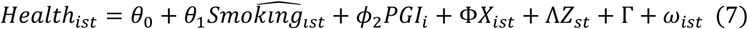

where *Health*_*ist*_ is the health outcome of interest, and *Smoking*_*ist*_ is the predicted value from the first stage. The coefficient of interest, *θ*_1_, is interpreted as the causal effect of smoking on health. The model includes the same vector of individual controls (*X*_*ist*_) and state-level controls (*Z*_*st*_), as the first stage. The term Γ represents the full set of fixed effects for survey year, birth year, adolescent state, and current state. Consistent with our primary analysis, these IV models are estimated on the pooled cross-sectional sample as per prior literature^8,10,31^.

## Data Availability

This study used restricted individual-level information from the HRS, and our contractual agreement does not permit public dissemination of the data. Details on how to access restricted data for the HRS can be found at https://hrs.isr.umich.edu/data-products/restricted-data. This study was approved by the Institutional Review Board (IRB) at the University of Wisconsin-Madison (2019-0924-CP003). HRS obtained informed consent from all participants.

## Disclosure Statement

The authors declare no conflicts of interest.

## Funding

Alemu received funding from the Neubauer Family Fellowship that covered the cost of attending his doctoral studies. Schmitz acknowledges funding from the National Institute on Aging (NIA): K99/R00 AG056599 and P30 AG017266. The content is solely the authors’ responsibility and does not necessarily represent the official views of the National Institutes of Health.

## Acknowledgments

We are grateful for the valuable feedback from Profs. William A. Masters, Steven A. Block, Cynthia Kinnan, Dan Benjamin, Patrick Turley, David Cesarini, Jason Fletcher, and Norma Coe. We thank seminar and conference participants at Tufts University, the “Frontiers in Genetics and Economics Conference hosted by BRIDGE (BRInging Data on Genetics to Economics), and lab meeting at the Social Science Genetic Association Consortium (SSGAC). We are thankful for the research assistance by Lucinda Toyoma.

## Notes

### Competing Interest Statement

The authors have declared no competing interest.

### Author Declarations

The Institutional Review Board (IRB) at the University of Wisconsin-Madison and the University of California Los Angeles (UCLA) gave ethical approval for this work.

