## Supplementary Information for "Exposure to Higher Cigarette Taxes during Adolescence Reduces Lifetime Smoking in Individuals with Elevated Genetic Risk"

### Notes

#### A. Correcting for mortality selection

To address potential bias from mortality selection, we employ an inverse probability weighting (IPW) approach. This method re-weights the estimation sample to make it more representative of the original full cohort, including those who did not survive until the genotyping wave. The weights are constructed based on the predicted probability of survival until 2006, the first year of genotyping in the HRS. We model this survival probability using the following probit model:

$$P(Alive_{isb} = 1) = \Phi(\alpha + \delta X_i + \lambda H_i + \gamma_b) \quad (8)$$

where the dependent variable is an indicator for whether respondent $i$ was alive in year 2006; $\Phi(.)$ is the cumulative distribution function of the standard normal distribution, which ensures the predicted probability lies between 0 and 1. The model includes a vector of baseline individual-level covariates, $X_i$, and a vector of baseline health indicators, $H_i$ (a quadratic in body mass index, and indicators for diabetes, heart problems, lung disease, cancer, stroke, self-reported health, and depression), along with birth year fixed effects, $\gamma_b$.

After estimating this model for the full initial sample, the inverse probability weight for each individual in our final estimation sample is calculated as the inverse of their predicted probability of survival. This procedure up-weights individuals who were observationally similar to those who did not survive, thereby correcting for potential selection bias.

#### B. Estimating travel distance as a proxy for tax avoidance behavior

To construct a proxy for tax avoidance behavior, we calculated the driving distance from each respondent's county of residence to the nearest state with a lower cigarette excise tax for each survey wave. This was accomplished using the Network Analyst extension in ArcGIS. We first obtained U.S. county and state boundary shapefiles and a national road network dataset from the U.S. Census Bureau. For each respondent in each survey wave, we identified their county of residence using restricted geographic data from the HRS and calculated its geographic centroid to serve as the origin point. For the destinations, we identified all neighboring states with a lower cigarette excise tax in that specific year and generated a series of points along their borders. Using the "Closest Facility" analysis tool, we then calculated the shortest driving route distance in miles along the road network from each origin (county centroid) to the nearest destination (point on a lower-tax state border). This wave-specific driving distance measure was then merged with our main dataset and included as a control variable in our regression models.

### 1. Supplementary Tables

**Table S1: Summary statistics of HRS study participants (1992-2016)**

| | (1) | (2) | (3) | (4) | (5) | (6) |
|---|---|---|---|---|---|---|
| **Panel A: Time invariant** | **Full Sample Mean (S.D.)** | **Current Smokers Mean (S.D.)** | **Never Smokers Mean (S.D.)** | **Former Smokers Mean (S.D.)** | **Difference (Current -Never) Mean (S.E.)** | **Difference (Former -Current) Mean (S.E.)** |
| Female (%) | 57.99 | 58.74 | 67.66 | 48.17 | -0.090*** (0.005) | -0.105***(0.005) |
| PGI Z-Score (Initiation) | -0.004(1.003) | 0.363(0.985) | -0.246(0.979) | 0.120(0.973) | 0.140***(0.010) | -0.243***(0.009) |
| PGI Z-Score (CPD) | 0.006 (1.00) | 0.12 (1.00) | -0.02 (0.99) | -0.005 (1.01) | 0.140*** (0.010) | -0.125*** (0.010) |
| PGI Z-Score (Cessation) | 0.006 (1.00) | -0.12 (0.96) | -0.002 (0.98) | 0.06 (1.03) | -0.119*** (0.009) | 0.176***(0.009) |
| **Panel B: Time variant** | | | | | | |
| Age (years) | 66.96 (11.01) | 61.45 (9.19) | 67.51 (11.39) | 68.21 (10.62) | -6.063*** (0.094) | 6.760***(0.092) |
| Education (years) | 13.19 (2.55) | 12.41 (2.39) | 13.43 (2.54) | 13.20 (2.67) | -1.017*** (0.023) | 0.785***(0.023) |
| Income (log) | 10.81(1.14) | 10.57(1.34) | 10.87(1.10) | 10.84(1.09) | -0.307***(0.013) | 0.270***(0.013) |
| Married (%) | 64.49 | 59.94 | 64.44 | 66.21 | -0.045*** (0.005) | 0.064***(0.005) |
| Ever smoked (%) | 57.02 | 100 | - | 100 | - | - |
| Currently smoking (%) | 13.73 | 100 | - | - | - | - |
| Former smoker (%) | 43.29 | - | - | 100 | - | - |
| Cigarettes per day | 3.53 (7.95) | 17.08 (8.40) | - | 15.47 (9.97) | - | - |
| Minimum age for cig. sale (years) | 18.05 (0.30) | 18.05 (0.28) | 18.05 (0.31) | 18.05 (0.29) | -0.006* (0.003) | 0.001(0.003) |
| Smoke-free air law (%) | 49.75 | 42.15 | 50.63 | 51.35 | -0.085*** (0.005) | 0.092***(0.005) |
| Cigarette tax during adolescence | 0.96 (0.17) | 0.94 (0.16) | 0.97 (0.18) | 0.96 (0.17) | -0.028*** (0.002) | 0.021***(0.002) |
| Cigarette tax while in HRS | 1.52 (0.63) | 0.86 (0.83) | 0.97 (0.86) | 0.99 (0.88) | -0.111*** (0.015) | 0.128***(0.008) |
| Travel distance to nearest lower tax state border (miles) | 48.93 (79.43) | 48.83 (77.18) | 46.20 (75.57) | 51.63 (83.59) | 2.631*** (0.736) | 2.804***(0.755) |
| Individuals (N) | 11,558 | 2,515 | 5,196 | 4,226 | 7,711 | |
| Person-wave observations | 104,651 | 14,347 | 44,910 | 42,654 | 59,257 | |

*Notes*: *** p<0.01, ** p<0.05, * p<0.1 with standard errors in parentheses. The coefficients in column 6 are from standard two-sample t-tests run assuming unequal variance of the covariates between current and never-smoker groups. The two-sample t-tests were run on the pooled sample, so the standard errors are adjusted for autocorrelation at the respondent level. All the coefficients in columns 2-5 represent the means and standard deviation of the variables. The PGI Z-scores are computed by subtracting the mean PGI from the raw PGI scores and dividing by the standard deviation.

**Table S2. Incremental R-squared of each covariate predicting smoking behavior outcomes**

| | Smoking Behavior Outcomes | | | |
|---|---|---|---|---|
| | **Ever smoker** | **Current smoker** | **Former smoker** | **CPD** |
| Smoking initiation PGI | 3.979 | 2.014 | - | - |
| Smoking cessation PGI | - | - | 0.556 | - |
| CPD PGI | - | - | - | 1.840 |
| Sex | 2.871 | 0.002 | 0.790 | 0.100 |
| Age (level, squared) | 0.280 | 3.934 | 6.710 | 3.290 |
| BMI (kg/meters$^2$) | 0.050 | 1.430 | 2.400 | 0.920 |
| Education (years) | 1.300 | 2.480 | 2.400 | 2.980 |
| Marital status | 0.460 | 0.960 | 1.000 | 0.660 |
| Income (quintiles) | 0.120 | 0.840 | 0.900 | 1.100 |
| Cigarette excise taxes | 0.070 | 0.000 | 0.000 | 0.500 |
| Other predictors | 0.050 | 0.000 | 0.100 | 0.000 |
| Total R-Squared | 9.130 | 11.660 | 14.756 | 11.390 |

*Notes*: This table shows the incremental R-squared of each covariate listed in the first column (expressed as a percent by multiplying by 100). The incremental R-squared of the PGIs is calculated by estimating the coefficient of determination ($R^2$) of the PGIs, after residualizing the effects of the first 20 principal components of the genetic relationship matrix. For the other covariates, the incremental R-squared is computed by taking the difference of $R^2$ between a base model that regresses the state-fixed effects on the smoking behavior outcome variables and subsequent models that sequentially add each predictor. Other predictors include a minimum age for sale, smoke-free air laws, and travel distance from the county of residence to the lower cigarette tax state border (level, squared, and cubed). The incremental R-squared values represent the share (%) of total variation each predictor explains.

**Table S3. Effects of cigarette taxes on current smoking by genetic risk profile, controlling for cigarette tax by PCs interactions**

| | |
|---|---|
| Adolescent Tax | -0.005** |
| | (0.024) |
| Adult Tax | -0.024*** |
| | (0.008) |
| Smoking Initiation PGI | 0.064*** |
| | (0.016) |
| Adolescent Tax x PGI | -0.026* |
| | (0.015) |
| Adult Tax x PGI | -0.001 |
| | (0.004) |
| Person-wave observations | 96,996 |
| Individuals (N) | 9,719 |
| R-squared | 0.110 |

*Notes*: This table presents coefficients from a linear probability model for current smoking status. The model is identical to the main specification (Table 1, Model 6) but includes interaction terms between each of the top 10 genetic principal components (PCs) and the adolescent and adult cigarette tax variables. These interactions help adjust for potential confounding due to ancestry-related structure that may correlate with environmental exposures, following Keller (2014). The dependent variable is an indicator for whether a respondent is a current smoker. Adolescent Tax is the average state cigarette excise tax experienced during adolescence, and Adult Tax is the average state tax experienced during older adulthood. PGI is the standardized polygenic index for smoking initiation. The model includes the full set of individual, state-level, and genetic controls, as well as all fixed effects detailed in the notes for Table 1. Robust standard errors, shown in parentheses, are two-way clustered at the individual and adolescent state-of-residence levels. *** p<0.01, ** p<0.05, * p<0.1.

**Table S4. Effects of smoking cessation on the likelihood of developing common chronic diseases and survival**

| | Lung Disease | | Heart Problem | | Any Cancer | | Diabetic | | Survival | |
|---|---|---|---|---|---|---|---|---|---|---|
| | (1) OLS | (2) 2SLS | (3) OLS | (4) 2SLS | (5) OLS | (6) OLS | (7) OLS | (8) 2SLS | (9) OLS | (10) 2SLS |
| Former Smoker | -0.074*** | -0.984*** | 0.011 | -1.222*** | 0.003 | -0.396* | 0.003 | -0.686*** | 0.018*** | 1.623*** |
| | (0.007) | (0.240) | (0.009) | (0.360) | (0.010) | (0.206) | (0.007) | (0.181) | (0.003) | (0.350) |
| F-stat (1st Stage) | - | 21.91 | - | 22.56 | - | 21.85 | - | 22.53 | - | 25.96 |
| Person-wave observations | 55,348 | 55,348 | 55,550 | 55,550 | 55,503 | 55,503 | 55,449 | 55,449 | 60,207 | 60,20 |
| Individuals (N) | 6,659 | 6,659 | 6,659 | 6,659 | 6,659 | 6,659 | 6,659 | 6,659 | 6,659 | 6,659 |

*Notes*: This table shows coefficients of naïve OLS (models 1, 3, 5, 7, and 9) and 2SLS (models 2, 4, 6, 8, and 10) regression models based on the specifications highlighted in equations (6-9). The dependent variables are indicators for whether respondents report ever being diagnosed with lung disease (models 1 and 2), heart problem (models 3 and 4), any cancer (models 5 and 6), diabetes (models 7 and 8), and survival at the time of survey (models 9 and 10). Models 1-8 were run on the HRS dataset that pools observations across all survey waves (1992-2016), conditional on survival at a given survey wave. The key predictor is an indicator variable for whether a respondent is a "Former Smoker". In the 2SLS models, smoking behavior or "Current Smoker" is instrumented by the average state cigarette excise tax that respondents were exposed during adolescence and while in the HRS. All regression models control for gender, BMI (level, squared), education (level, squared), marital status, household income, state, birth year, and survey-year fixed effects. Robust standard errors clustered at the individual and state of residence levels are shown in parenthesis. *** p<0.01, ** p<0.05, * p<0.1.

**Table S5. Effects of smoking intensity (CPD) on the likelihood of developing common chronic diseases and survival**

| | Lung Disease | | Heart Problem | | Any Cancer | | Diabetic | | Survival | |
|---|---|---|---|---|---|---|---|---|---|---|
| | (1) | (2) | (3) | (4) | (5) | (6) | (7) | (8) | (9) | (10) |
| | OLS | 2SLS | OLS | 2SLS | OLS | OLS | OLS | 2SLS | OLS | 2SLS |
| CPD | 0.005*** | 0.032*** | 0.001 | 0.022*** | 0.001 | 0.018*** | 0.001* | 0.014*** | -0.001*** | -0.035*** |
| | 0.001 | (0.009) | (0.001) | (0.006) | (0.001) | (0.006) | (0.001) | (0.005) | (0.000) | (0.008) |
| F-stat (1st Stage) | - | 16.10 | - | 15.93 | - | 16.73 | - | 16.14 | - | 21.43 |
| Person-wave observations | 20,475 | 20,475 | 20,615 | 20,615 | 20,569 | 20,569 | 20,559 | 20,559 | 20,564 | 20,564 |
| Individuals (N) | 2,563 | 2,634 | 2,634 | 2,634 | 2,634 | 2,634 | 2,634 | 2,634 | 2,634 | 2,634 |

*Notes*: This table shows coefficients of naïve OLS (models 1, 3, 5, 7, and 9) and 2SLS (models 2, 4, 6, 8, and 10) regression models based on the specifications highlighted in equations (6-9). The dependent variables are indicators for whether respondents report ever being diagnosed with lung disease (models 1 and 2), heart problem (models 3 and 4), any cancer (models 5 and 6), diabetes (models 7 and 8), and survival at the time of survey (models 9 and 10). All models were run on the HRS dataset that pools observations across all survey waves (1992-2016), conditional on being a current smoker, while models 1-8 were further restricted to those who were alive in the specific wave under consideration. The key predictor is "CPD" representing the average number of cigarettes respondents (current smokers) reported smoking daily. In the 2SLS models, smoking behavior or "cigarettes smoked per day (CPD)" is instrumented by the average state cigarette excise tax that respondents were exposed to while in the HRS. All regression models control for gender, BMI (level, squared), education (level, squared), marital status, household income, state, birth year, and survey-year fixed effects. Robust standard errors clustered at the individual and state of residence levels are shown in parentheses. *** p<0.01, ** p<0.05, * p<0.1.

**Figure S1. Schematic showing derivation of the estimation sample**

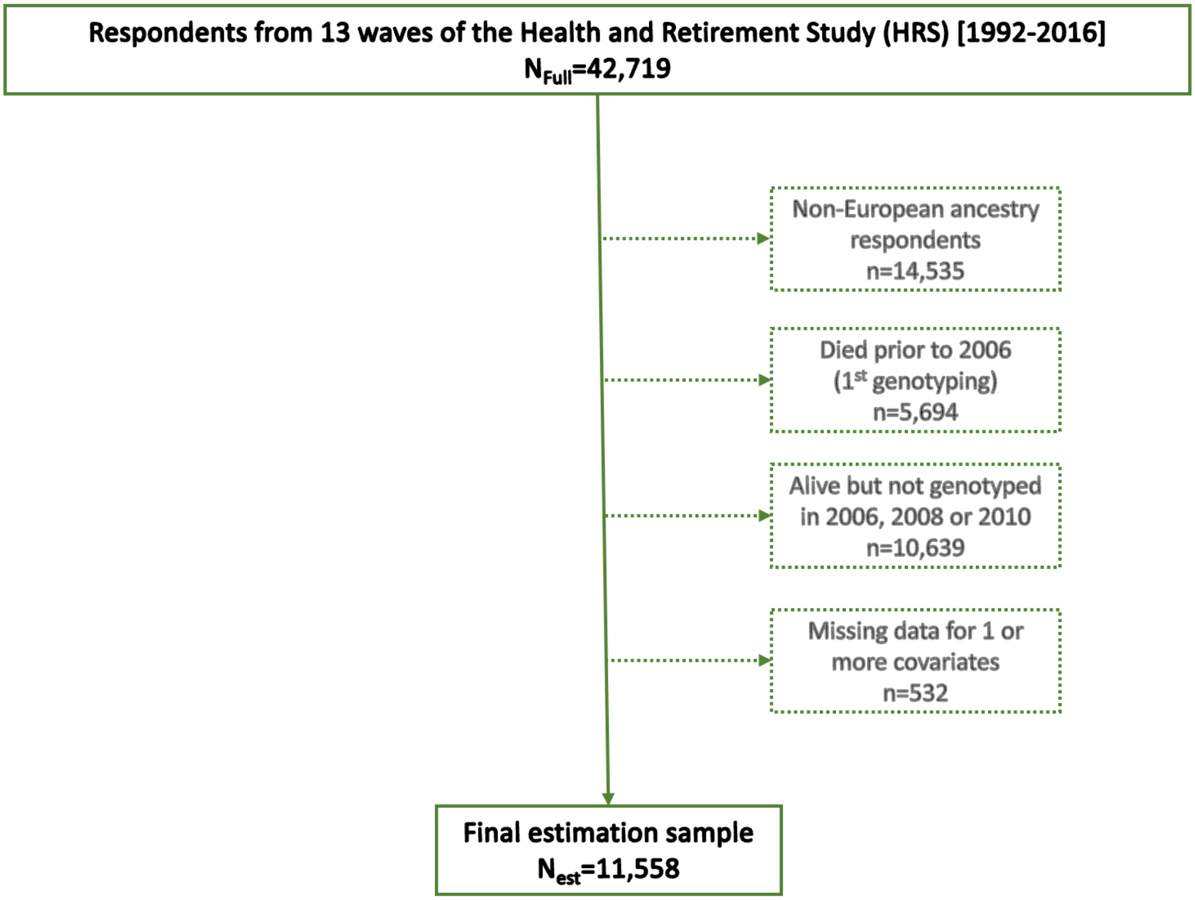


*Notes*: This schematic shows the steps followed to reach the estimation sample. The dashed rectangular boxes show the number of individuals excluded from our estimation sample. All non-European ancestry respondents were first excluded as PGIs developed using predominantly European-ancestry samples do not adequately predict phenotypes in non-European samples. The sample is further restricted by removing European-ancestry respondents who were deceased before the first genotyping (2006) and those who were alive in 2006 but were not genotyped. Finally, respondents with missing values for one or more regression model covariates were excluded, with most exclusions driven by missing adolescent state-of-residence information needed to construct cigarette tax exposure histories, yielding a final estimation sample of 11,558.

**Figure S2. State tobacco excise taxes by year**

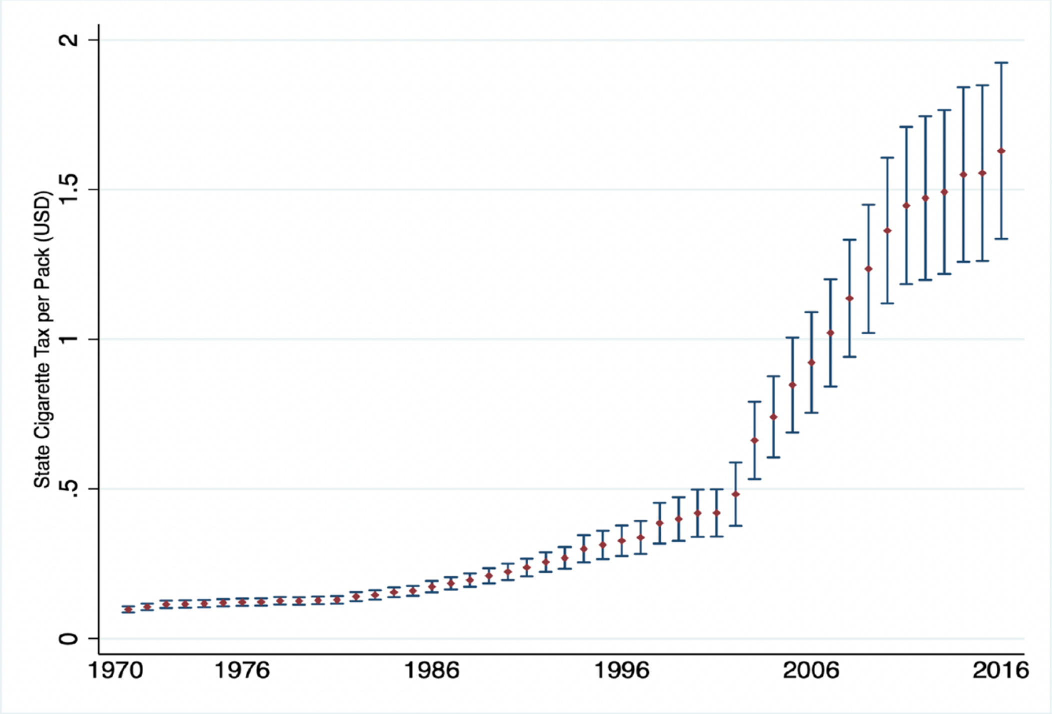


**Note:** Red dots represent average tobacco excise taxes for each year and arrows show the respective 95 percent confidence interval. **Source**: Orzechowski & Walker (2016). Tax burden on tobacco: The historical compilation, vol. 51.

**Figure S3. Spatial variation in state tobacco excise taxe across states in 2016**

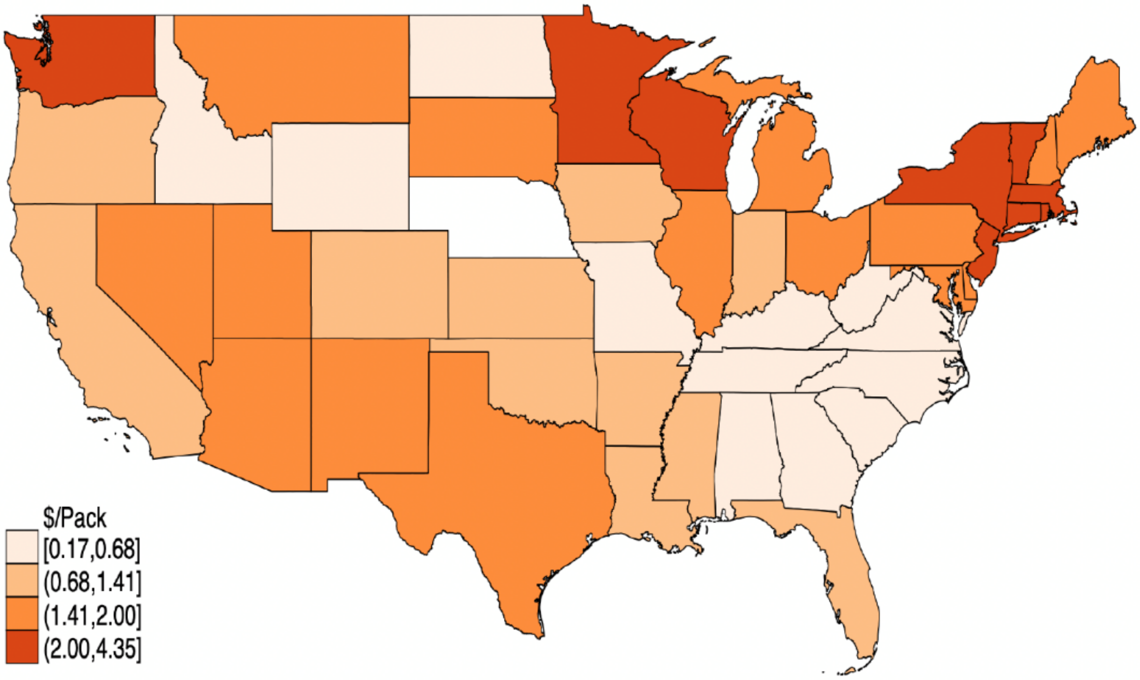


**Source**: Orzechowski & Walker (2016). Tax burden on tobacco: The historical compilation, vol. 51.

**Figure S4. Predicted probability of becoming a former smoker by PGI and life course timing of cigarette excise taxes**

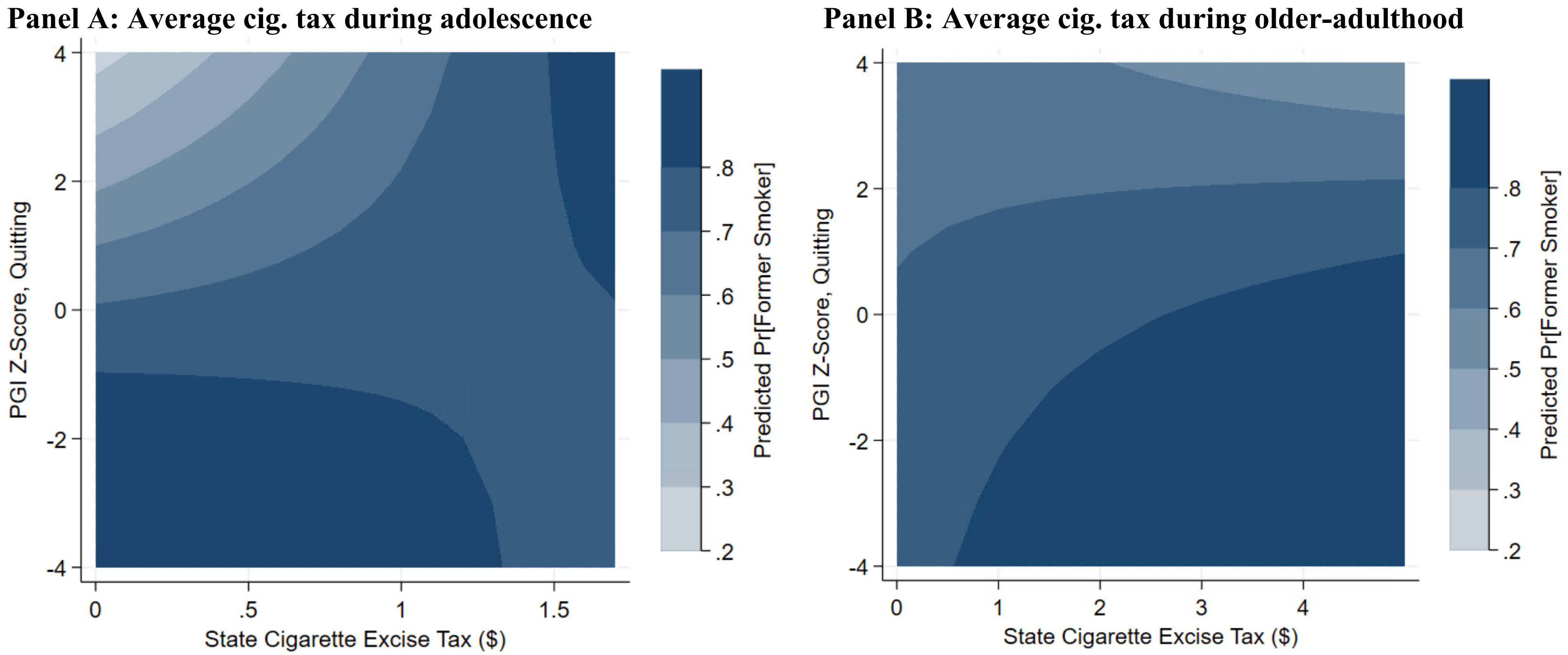


*Notes*: These contour plots show respondents' likelihood of quitting smoking by average cigarette excise tax during adolescence (panel A) and while in the HRS (panel B) by smoking intensity PGI for ever-smokers. The 3rd axis (z-axis) of the contour plot depicts the marginal predicted probabilities, which were estimated using a logit regression model run on the HRS dataset that pools observations across all survey waves as per equation (4). Contours of similar color represent identical predicted probabilities of being a lifetime smoker.

**Figure S5. Predicted number of cigarettes smoked per day by PGI and life course timing of cigarette excise taxes**

**Panel A: Average cig. tax during adolescence**

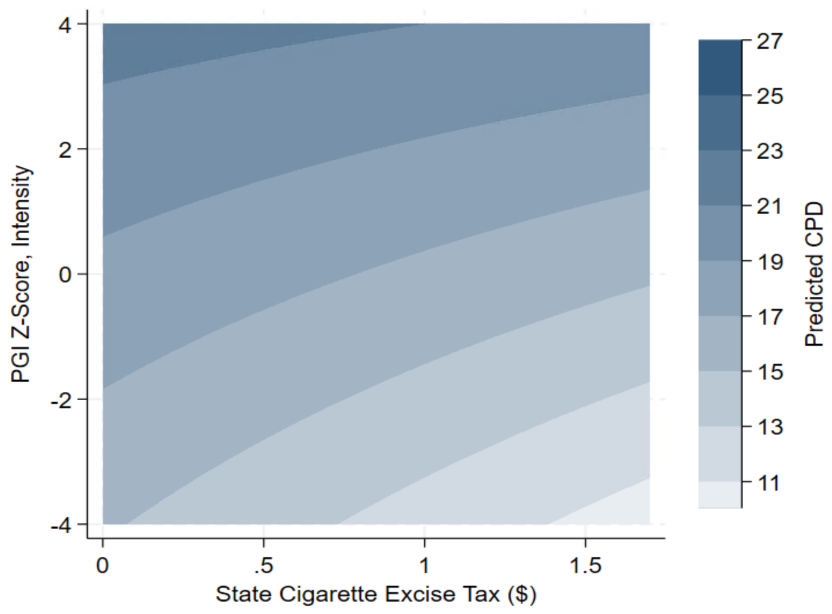


**Panel B: Average cig. tax during older-adulthood**

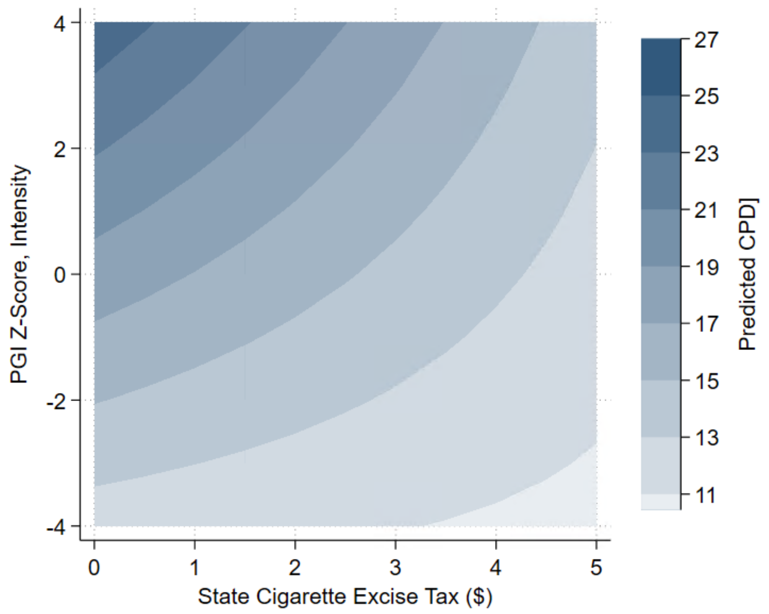


*Notes*: These contour plots show the predicted number of cigarettes smoked per day (CPD) for respondents who experienced different levels of average cigarette excise tax during their adolescence (panel A) and while in the HRS (panel B) by the genetic predisposition to smoking intensity (PGI Z-score), conditional on being a current smoker. The 3$^{rd}$ axis (z-axis) of the contour plot depicts the marginal predicted CPDs, and were estimated using an OLS regression model run on the HRS dataset that pools observations across all survey waves as per equation (4). Contours of similar color represent identical predicted probabilities of being a lifetime smoker.
